# Clinical Impact of Contouring Variability for Prostate Cancer Tumor Boost

**DOI:** 10.1101/2024.01.29.24301942

**Authors:** Allison Y. Zhong, Asona J. Lui, Svetlana Kuznetsova, Karoline Kallis, Christopher Conlin, Deondre D. Do, Mariluz Rojo Domingo, Ryan Manger, Patricia Hua, Roshan Karunamuni, Joshua Kuperman, Anders M. Dale, Rebecca Rakow-Penner, Michael E. Hahn, Uulke A. van der Heide, Xenia Ray, Tyler M. Seibert

## Abstract

**Purpose:** The focal radiotherapy (RT) boost technique was shown in the FLAME trial to improve prostate cancer outcomes without increasing toxicity. This technique relies on the accurate delineation of prostate tumors on MRI. The ReIGNITE RT Boost study evaluated radiation oncologists’ accuracy when asked to delineate prostate tumors on MRI and demonstrated high variability in tumor contours. We sought to evaluate the impact of contour variability and inaccuracy on predicted clinical outcomes. We hypothesized that radiation oncologists’ contour inaccuracies would yield meaningfully worse clinical outcomes.

**Materials & Methods:** 44 radiation oncologists and 2 expert radiologists contoured prostate tumors on 30 patient cases. Of these cases, those with CT simulation or diagnostic CT available were selected for analysis. A knowledge-based planning model was developed to generate focal RT boost plans for each contour per the FLAME trial protocol. Probability of biochemical failure (BF) was determined using a model from the FLAME trial. The primary metric evaluated was delta BF (ΔBF *=* Participant BF – Expert BF). An absolute increase in BF ≥5% was considered clinically meaningful.

**Results:** 8 patient cases and 394 target volumes for focal RT boost planning were included in this analysis. In general, participant plans were associated with worse predicted clinical outcomes compared to the expert plan, with an average absolute increase in BF of 4.3%. 37% of participant plans were noted to have an absolute increase in BF of 5% or more.

**Conclusion:** Radiation oncologists’ attempts to contour tumor targets for focal RT boost are frequently inaccurate enough to yield meaningfully inferior clinical outcomes for patients.

## Introduction

In the FLAME phase III randomized controlled trial, adding a focal radiotherapy (RT) boost to prostate tumors visible on multiparametric MRI improved disease-free and regional/distant metastasis-free survival for patients with intermediate- and high-risk prostate cancer, without increasing toxicity [1,2]. This trial demonstrated that the probability a patient would experience biochemical failure (cancer recurrence) was predicted by the RT dose delivered to the visible tumor. Higher tumor doses yielded better treatment outcomes, and lower tumor doses yielded more treatment failures.

The focal RT boost technique relies on the accurate delineation of prostate tumors on MRI. A recent prospective study, ReIGNITE RT Boost, evaluated radiation oncologists’ accuracy when asked to delineate prostate tumors on MRI [3]. High variability was observed in tumor contours. When using conventional MRI alone, participants completely missed the tumor (zero overlap with the true target) in a median of 13.6% of attempts (IQR 9.1-23.6%). Accuracy and reliability of tumor contours improved considerably when participants were given advanced diffusion MRI maps (called the Restriction Spectrum Imaging restriction score, or RSIrs) that improve prostate cancer conspicuity. Complete misses, for example, were much more uncommon when using RSIrs (median 0.0%, IQR 0.0% - 4.3%). Nonetheless, contours still varied considerably between radiation oncologists. Given smooth radiation dose plans and RT penumbra, it is unclear how near misses and partial overlap will affect patient outcomes.

Here, we evaluate the impact of contour variability and inaccuracy on predicted clinical outcomes. We measured the RT dose delivered to the true tumor if a ReIGNITE participant’s contour were used to generate the focal RT boost plan instead of using the guidance of an expert radiologist, as was done in the FLAME trial. Using the model from the FLAME trial [2], we then calculated the probability of biochemical failure for each RT plan. We hypothesized that radiation oncologists’ contour inaccuracies would yield meaningfully worse clinical outcomes.

## Methods & Materials

### ReIGNITE Study

44 radiation oncologists with a range of experience treating prostate cancer were enrolled as participants. All study recruitment materials, communications, and procedures were approved by the Institutional Review Board (IRB). Participants were asked to contour tumors on 20 patient cases of clinically localized intermediate- or high-risk prostate cancer in each of two sessions at least 1 month apart. In each case, participants were provided either conventional MRI alone or conventional MRI with RSIrs overlay. 10 cases were repeated between the two sessions, with RSIrs either added or removed; the participants were not made aware of this. Provided MRI sequences included T2-weighted, ADC, and DWI (b=0 and b=2000 s/mm^2^). In addition, articipants were given clinical information regarding each case: including the patient’s age, PSA at time of MRI, number of positive biopsy cores, location of positive biopsy cores, and radiologic description of the tumor. Contouring was performed on the MIM Zero Footprint™ (ZFP) platform (MIM Software, Cleveland, OH).

Expert-defined lesions were created by the consensus of a radiation oncologist (with three years of experience) and two board-certified, subspecialist genitourinary radiologists (with five years and seven years of experience, respectively). This was done for all patient cases on conventional MRI alone, as the clinical standard per the FLAME trial is to target the tumor visible on MRI.

For this present study, we included all patient cases from ReIGNITE for which either a CT simulation or a diagnostic CT was available.

### OAR and Target Volume Definitions

Where a CT simulation was available, organs at risk (OARs) had been contoured per clinical routine. For those with only a diagnostic CT, OARs were contoured manually. T2-weighted MRI for each case was rigidly co-registered to the CT to optimize registration specifically of the prostate, and the corresponding transformation was applied to the expert-defined tumor target, as well as to each participant’s target contour.

### Knowledge-based RT Planning

To ensure RT planning was unbiased, we developed a knowledge-based planning (KBP) automated algorithm to generate RT plans with focal tumor boost per the FLAME trial protocol: 77 Gy in 35 fractions to the whole prostate and an integrated boost up to 95 Gy to the focal target, provided no normal tissue constraints were violated. To facilitate the development of the KBP algorithm, we used Varian RapidPlan (Varian Medical Systems, Palo Alto, USA), a vendor-based solution for knowledge based planning [4] that is widely used in the field [5–10]. A total of 62 clinical plans with contoured targets (prostate and boosted lesion) and OARs (bladder, rectum, bowel, sigmoid, urethra, penile bulb, right femur, and left femur) were added to the training set for the model. RapidPlan then uses geometrical features extracted between the set of targets and OARs and the OAR dose-volume histograms (DVH) to build models that can predict the achievable OAR DVHs for a new patient. Additionally, the set of OAR DVH models is linked to a template of weighted optimization metrics that use the patient-specific predicted dose values to optimize and calculate dose for new plans.

This allows for fully automated, high-quality planning. The plan optimization metrics for our model (**Table 1**) reflect those used in the FLAME trial as well as an additional urethra constraint proposed in a subsequent normal tissue complication probability analysis of FLAME participants [11].

**Table 1.** Knowledge-based planning (KBP) optimization metrics.

| Structure | Limit | Vol (%) | Dose (cGy) | Priority |
| --- | --- | --- | --- | --- |
| **GTV_xxx** | Upper | 0.0 | 9394 | 80 |
|  | Lower | 100.0 | 9548 | 70 |
| **PTV78_DVH_xxx** | Lower | 100.0 | 7854 | 200 |
| **PTV78_Opt_xxx** | Upper | 0.0 | 7931 | 200 |
|  | Lower | 100.0 | 7854 | 200 |
| **Bladder** | Upper | 1.1 | 7315 | 100 |
|  | Upper | 3.7 | 3850 | 100 |
|  | Upper | 8.5 | 1925 | 30 |
|  | Upper | 0.0 | 7700 | 125 |
|  | Line |  |  | 50 |
| **Femur L** | Line |  |  | 50 |
| **Femur R** | Line |  |  | 50 |
| **Penile Bulb** | Line |  |  | 50 |
| **Rectum** | Upper | 4.3 | 7315 | 100 |
|  | Upper | 10.6 | 5775 | 90 |
|  | Upper | 19.9 | 3850 | 90 |
|  | Upper | 30.6 | 2695 | 90 |
|  | Upper | 0.0 | 7700 | 125 |
|  | Line |  |  | 50 |
| **Sigmoid** | Upper | 0.0 | 5775 | 70 |
|  | Upper | 0.0 | 3850 | 70 |
|  | Upper | 0.0 | 5159 | 125 |
| **Urethra** | Upper | 0.0 | 8085 | 125 |

We tested the KBP model on 10 unrelated patient cases from our institution that were treated with focal RT boost. A dosimetrist manually generated optimal plans using the FLAME protocol for each of the 10 cases, using the focal tumor target actually treated for that patient. KBP plans were generated for the 10 cases, and the resulting dose metrics and 3D dose distributions were compared to the clinical plan used for those patients to validate the model (**Table 2**). To further validate the KBP model, we confirmed that it gave expected results for focal RT plans using the expert contours (**Table 3**). We then applied the algorithm to each participant’s tumor contour and compared dosimetric parameters to those achieved when using the expert-defined tumor.

**Table 2.** Knowledge-based planning (KBP) model validation with an independent set.

| Parameter | Dose Constraint | Mean±Std |
| --- | --- | --- |
| Prostate V100% | $V100\% \geq 94-95\%$ | 94.84±0.30 % |
| Rectum D<sub>1cc</sub> | $D_{1cc} \leq 77 \text{ Gy}$ | 78.29±0.27 Gy |
| Bladder D<sub>1cc</sub> | $D_{1cc} \leq 80 \text{ Gy}$ | 79.33±1.27 Gy |
| Urethra D<sub>0.1cc</sub> | $D_{0.1cc} \leq 80 \text{ Gy}$ | 79.91±1.05 Gy |
| Lesion D98 | $D98 > 95 \text{ Gy}$ | 85.42±3.00 Gy |
| Lesion D98 | $D98 > 120\% \text{ Rx}$ | 110.93±3.89%Rx |

**Table 3.** Results of knowledge-based planning (KBP) model on validation plans. The generated validation plans were based on expert contours. Clinical plans were generated by an experienced dosimetrist using clinical target and organ-at-risk volumes. All plans were normalized to meet the Prostate V100% metric, leading to no difference between Clinical and KBP plans. Other parameters were allowed to vary.

| Parameter | Dose Constraint | Clinical Mean±Std | KBP Mean±Std | Difference (KBP-Clinical) |
| --- | --- | --- | --- | --- |
| Prostate V100% | V100% ≥ 94-95% | 95±0% | 95±0% | 0% |
| Rectum D<sub>1cc</sub> | D<sub>1cc</sub> ≤ 103-105 % | 100.46±1.10% | 100.85±0.99% | -0.39±1.74% |
| Bladder D<sub>1cc</sub> | D<sub>1cc</sub> ≤ 105% | 101.56±0.34% | 102.44±0.35% | -0.88±0.66% |
| Urethra D<sub>0.1cc</sub> | D<sub>0.1cc</sub> ≤ 105-107 | 101.88±0.51% | 102.33±0.34% | -0.50±0.80% |
| Lesion Expert D98 | D98 > 120% Rx | 113.90±1.52% Rx | 114.78±7.57%Rx | -0.88±6.10% |

### RT Plan Evaluation & Statistical Tests

For each plan, we calculated the dose covering 98% of the expert-defined tumor (D98), which was associated with probability of biochemical failure (BF) at 7 years in a model from the FLAME trial (**Figure 1**). The primary metric we evaluated was delta BF probability (ΔBF), which was calculated as (Participant BF – Expert BF). A contour that led to a ≥5% increase in BF was considered clinically meaningful. The percentage of participants whose contours yielded a clinically meaningful BF increase was also calculated for each case. Secondary metrics included D98, BF, delta D98 (ΔD98 = Participant D98 – Expert D98), and the OAR dose metrics.

**Figure 1.**
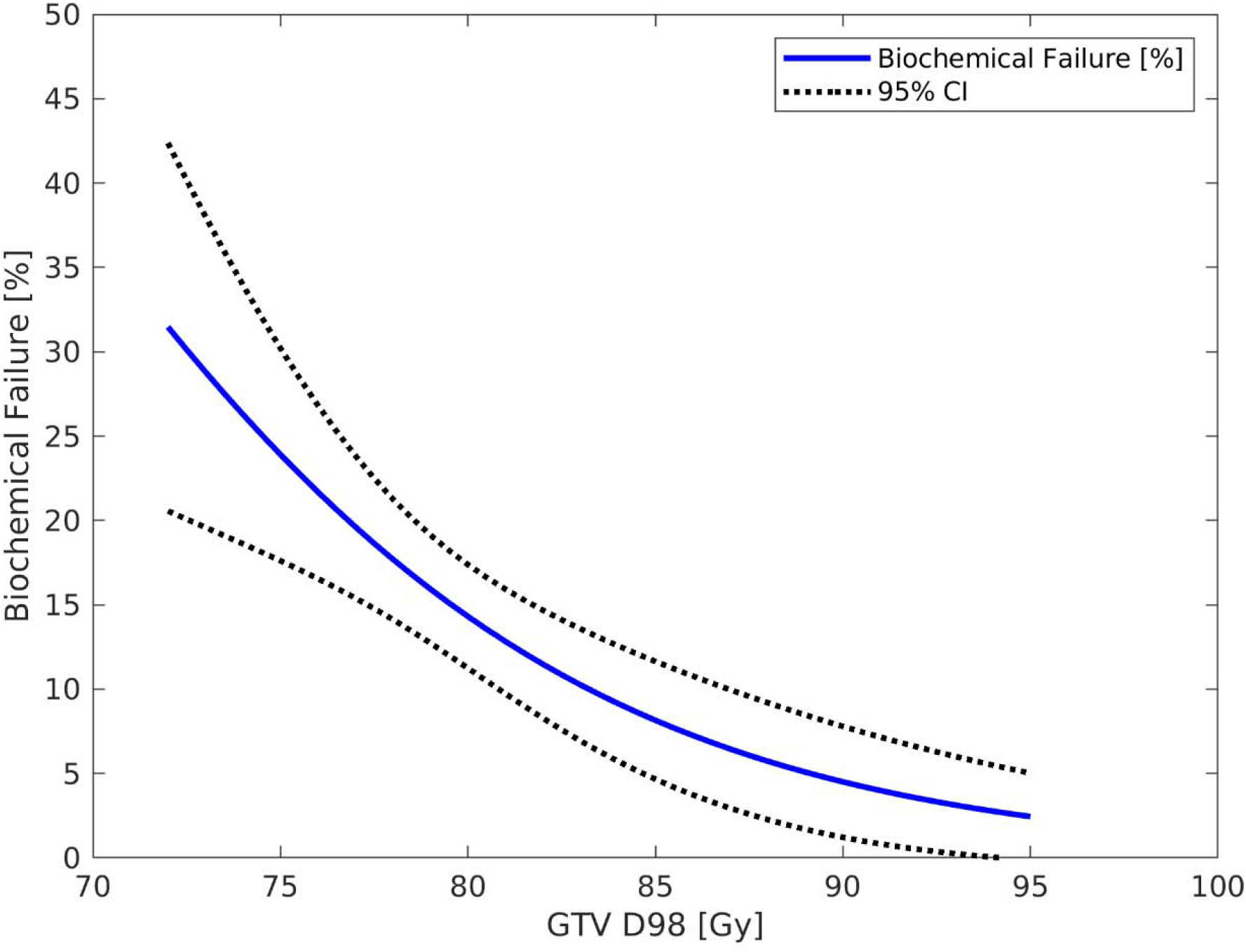
Biochemical failure model used in the FLAME trial [2].

Case-wise t-tests were performed for ΔBF and ΔD98 for all patient cases. For cases that were contoured both with and without RSIrs, paired t-tests were performed to compare ΔBF and ΔD98 with and without RSIrs.

## Results

8 patient cases from the ReIGNITE study had available pelvic CT scans and were included for analysis (**Table 4**). 2 cases had CT simulation available, while 6 cases had diagnostic CT only. 3 cases were contoured on conventional MRI only, 2 cases were contoured on conventional MRI + RSIrs only, and 3 cases were contoured on both with and without RSIrs. In total, 394 participant target volumes and 8 expert target volumes were used to generate RT plans. All plans had adequate coverage of the prostate and met all key normal tissue constraints.

**Table 4.** Characteristics of 8 patient cases included in this study.

|  |  |  |
| --- | --- | --- |
| **Median age, in years (IQR)** |  | 70 (67 - 78) |
| **Median PSA at time of MRI, in ng/mL (IQR)** |  | 8.7 (7.7 – 18.8) |
| **Clinical stage** | T1c | 6 |
|  | T2a | 1 |
|  | T2c | 1 |
| **PI-RADS v2.1 score per lesion** | 3 | 2 |
|  | 4 | 3 |
|  | 5 | 3 |
| **Biopsy status** | Systematic | 3 |
|  | Systematic & targeted | 5 |
| **Worst Gleason score per lesion** | 3+4 | 4 |
|  | 4+3 | 2 |
|  | 4+4 | 1 |
|  | 4+5 | 1 |
| **Underwent prostatectomy** |  | 2 |
| **Pathological stage** | T2a | 1 |
|  | T3b | 1 |

In general, using radiation oncologist participants’ target volumes yielded worse BF and D98 values compared to the expert target volume (**Figure 2**). Across all participants’ plans, BF increased by an average of 4.3%. A clinically meaningful absolute increase in BF of ≥5% was seen in 37% of the participant plans.

**Figure 2.**
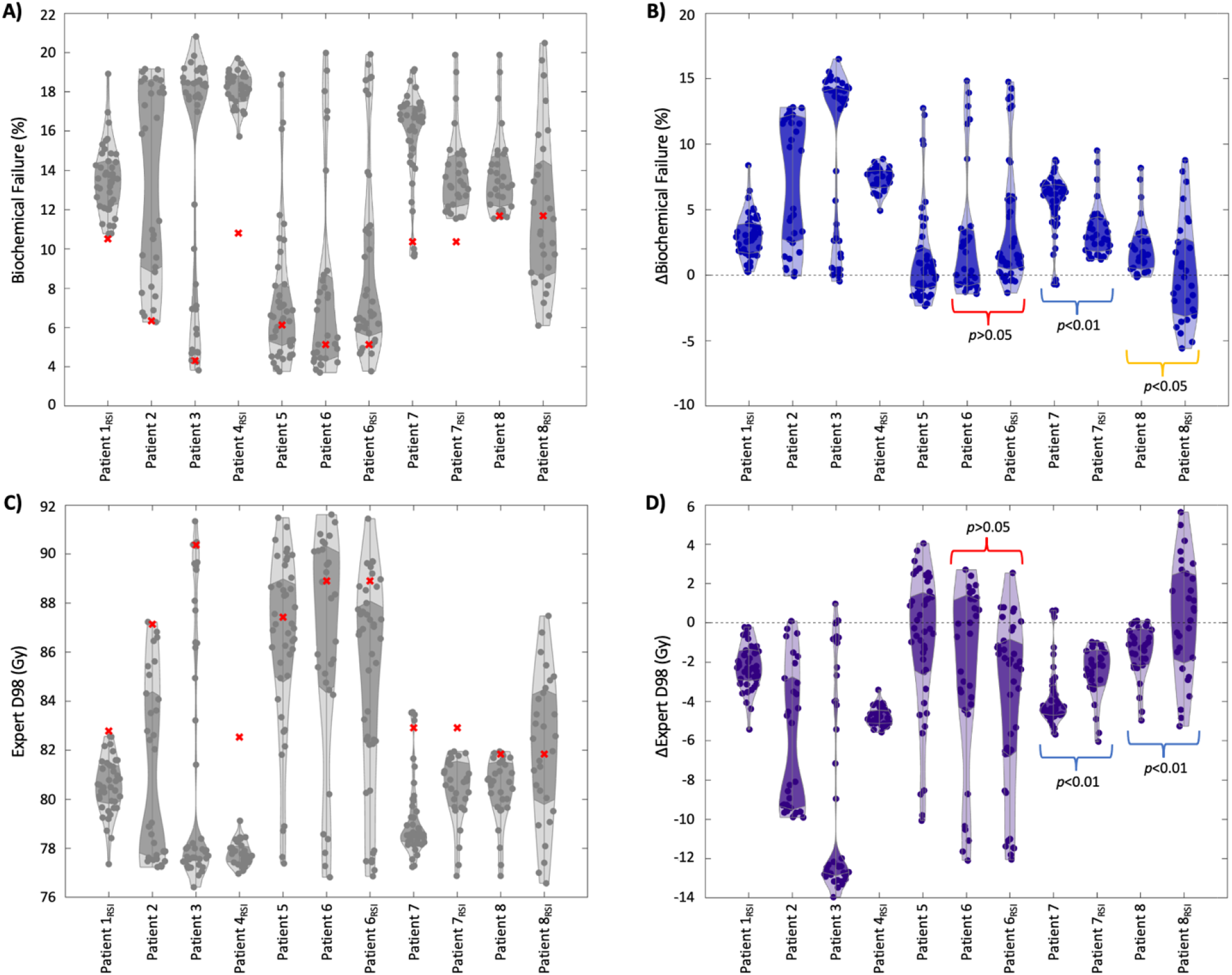
Violin plots for 8 patient cases for A) biochemical failure (BF), B) ΔBF = Participant BF – Expert BF, C) D98 of expert-defined tumor, and D) ΔD98 = Participant D98 – Expert D98. Predicted outcomes for the plans based on the expert contours are shown in red.

Case-wise t-tests comparing participant plans to expert plans demonstrated a statistically significant difference for ΔBF and ΔD98 in 6 of 8 patient cases (*p*<0.01) (**Table 5**). Among the 3 cases that had contours with and without RSI (Patients 6-8), paired t-tests showed a statistically significant improvement in both ΔBF and ΔD98 with RSIrs for Patients 7 and 8.

**Table 5.**
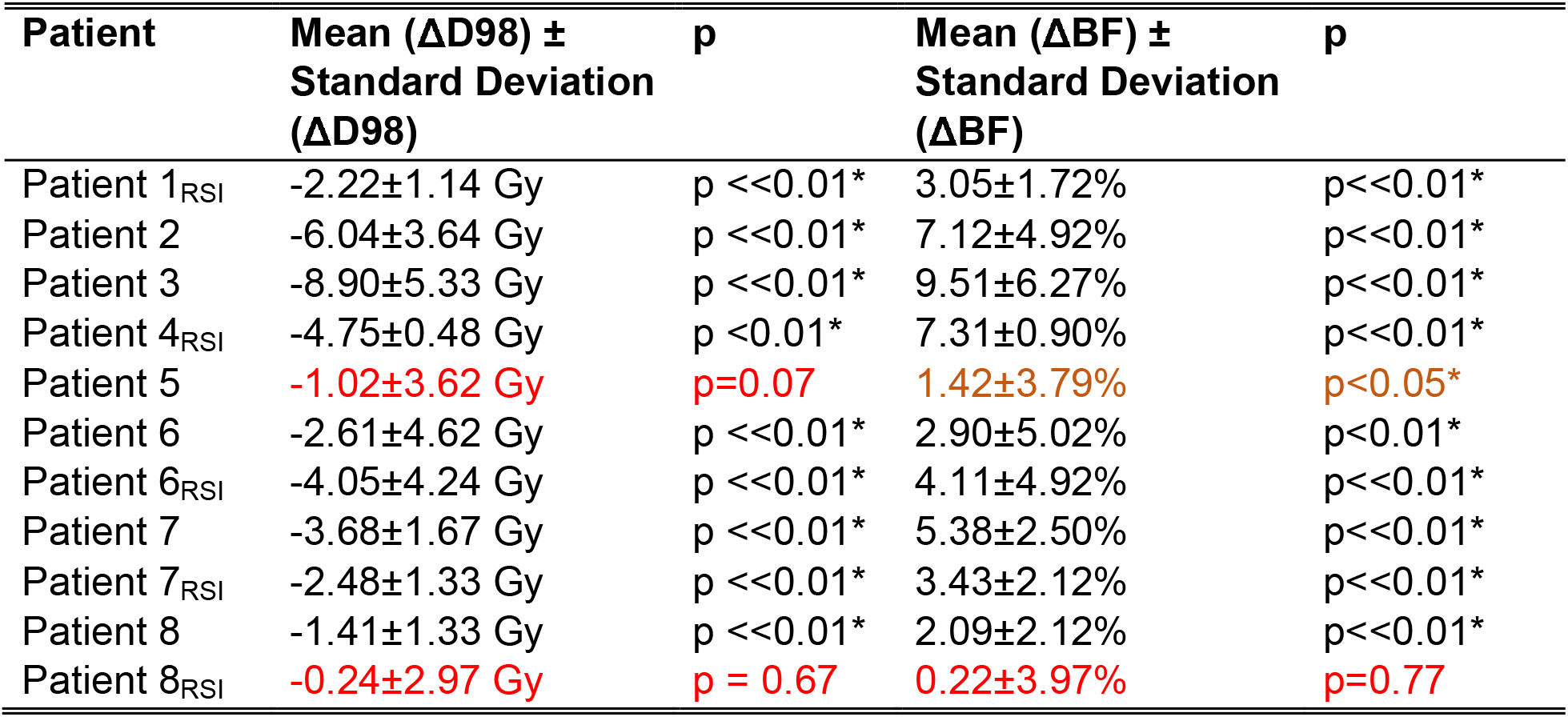
Case-wise t-tests of ΔD98 = Participant D98 – Expert D98 and ΔBF = Participant BF – Expert BF. *Significant with p<0.05.

## Discussion

Overall, using radiation oncologist participants’ target volumes yielded plans with worse predicted clinical outcomes compared to the expert target volume. On average, participants’ target volumes led to a predicted 4.3% absolute increase in BF. In more than one-third of participant plans (37%), the absolute increase in BF exceeded our *a priori* threshold for clinical significance (≥5% absolute increase in BF). Thus, the high tumor contour variability demonstrated in the ReIGNITE study is expected to adversely impact clinical outcomes for patients.

ReIGNITE showed that one way to improve radiation oncologists’ contouring accuracy is to provide RSIrs maps [3]. The present study was not powered for full evaluation of the impact of RSIrs on treatment plans, with only 3 cases included. Nonetheless, of the 3 cases that allowed for comparison of contours with and without RSIrs, 2 cases showed a statistically significant improvement in ΔBF and ΔD98 with RSIrs. Other strategies to improve contouring accuracy for MRI-visible prostate tumors include involvement of expert radiologists in target delineation and dedicated training for radiation oncologists [12,13]. Alternative imaging modalities like PSMA PET may also prove useful [14–18]. Automated target delineation using artificial intelligence has potential, though these currently still require considerable expert oversight [19,20]. For less experienced or resourced settings, one could consider adding a margin around the gross tumor volume to account for contouring uncertainties. Future studies should measure both the feasibility of these strategies and their impact on predicted clinical outcomes of focal RT boost. Meanwhile, it is nonetheless noted that all calculated BFs in the present study were lower than that if the prostate were treated only with a uniform dose of 77 Gy, illustrating that even somewhat inaccurate contours may still benefit the patient.

In some instances, an imprecise contour led to a higher dose than that seen in the expert plan. This was especially prevalent for Patients 5, 6, and 8_RSI_. Upon inspection of these cases, it appeared that participants tended to draw their contours larger than the expert-defined lesion, while encompassing the true target (**Figure 3**), which likely spread out the high dose away from OARs. In clinical practice, a dosimetrist would recognize the potential for improving a plan with the correct target, e.g., by inserting an optimization structure to better target the lesion. This indicates that the KBP model used in this study has room for further improvement to achieve more optimal planning.

**Figure 3.**
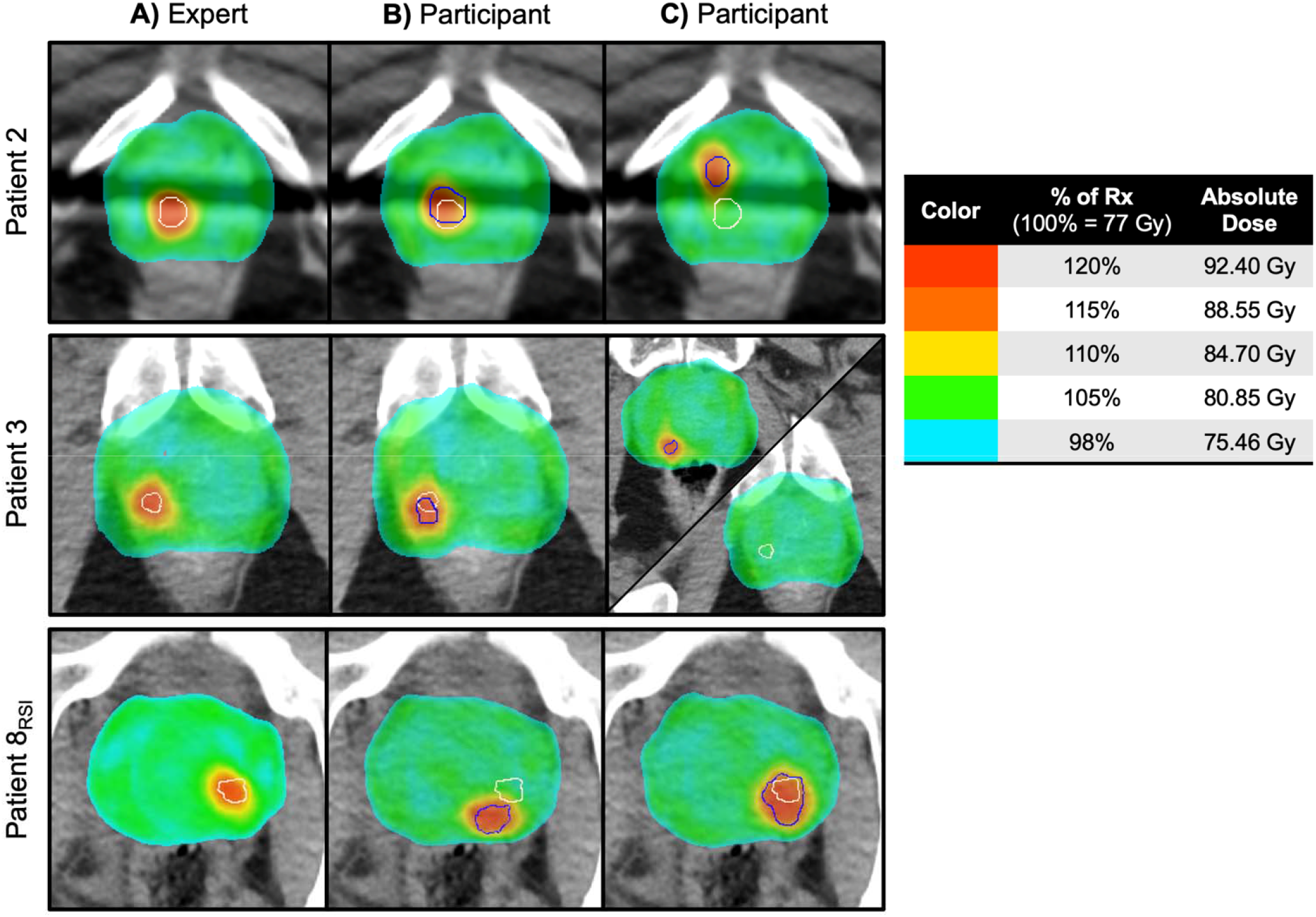
Example contours with dose overlays for 3 patient cases: A) Expert, B-C) Participants. Expert contours are shown in white, and participant contours are shown in blue. The greater the accuracy of the participant contour, the higher the dose delivered to the true tumor. When the participant drew the contour larger than and including the expert contour (such as in Patient 8(C)), this led to a higher dose to the true tumor than that seen in the expert plan.

The present study inherits some limitations from the ReIGNITE study that serves as the source of data. Images used in ReIGNITE were from a single institution, which may limit generalizability. Radiation oncologist participants, on the other hand, were from 9 countries and many institutions. Additionally, only 8 of the 30 patient cases from ReIGNITE were analyzed here. Still, we observed statistically and clinically significant results, suggesting realistic variability in target volumes can impact patient outcomes.

## Conclusion

Radiation oncologists’ attempts to contour tumor targets for focal RT boost are frequently inaccurate enough to yield meaningfully inferior clinical outcomes for patients. Further studies are warranted to investigate the impact of RSIrs on predicted clinical outcomes of focal RT boost.

## Data Availability

De-identified data are available to bona fide researchers for non-commercial use upon request.

## Notes

### Competing Interest Statement

AJL reports consulting for MIM Software. AMD is a Founder of and holds equity in CorTechs Labs, Inc, and serves on its Scientific Advisory Board. He is a member of the Scientific Advisory Board of Human Longevity, Inc. and receives funding through research agreements with GE Healthcare. RRP has an equity interest in CorTech Labs and Curemetrix, serves on the Scientific Advisory Board of Imagine Scientific, and receives research funding from GE Healthcare. MEH reports honoraria from Multimodal Imaging Services Corporation and research funding from GE Healthcare. UAvdH reports research funding from the Dutch Cancer Society and travel support from Elekta AB. XR reports consulting for KM Pharmaceutical Consulting LLC, receives research funding from Varian Medical Systems and Siemens Healthineers, and serves as vice chair of AAPM TG-395. TMS reports honoraria from Varian Medical Systems, WebMD, GE Healthcare, and Janssen; he has an equity interest in CorTechs Labs, Inc. and serves on its Scientific Advisory Board; he receives research funding from GE Healthcare through the University of California San Diego. These companies might potentially benefit from the research results. The terms of this arrangement have been reviewed and approved by the University of California San Diego in accordance with its conflict-of-interest policies.

### Funding Statement

This work was supported, in part, by the National Institutes of Health (NIH/NIBIB K08 EB026503, NIH/NCI U54CA132384, U54CA132379, UL1TR001442), the American Society for Radiation Oncology (ASTRO), the Prostate Cancer Foundation, the Radiological Society of North America (RSNA), the American College of Radiation Oncology (ACRO), the Grillo-Marxuach Family Fellowship at the Moores Cancer Center of UC San Diego, and the US Department of Defense (CDMRP PC220278).

### Author Declarations

The Institutional Review Board of UC San Diego gave ethical approval for this work.

